# Feasibility, usability, acceptability and Efficacy of a novel leg strengthening device (S-Press) for strengthening leg muscles and improving physical impairment during hospital admission

**DOI:** 10.1101/2024.01.15.24301211

**Authors:** TM. Maden-Wilkinson, C. Griffiths, B. Lakkappa, K. Walker, CL Kennerley, JJ. Turner

**Affiliations:** Physical Activity, Wellness and Public Health Research Group, Advanced Wellbeing Research Centre, Health Research Institute, Sheffield Hallam University, Sheffield, UK; Northamptonshire Healthcare NHS Foundation Trust, Bevan House Kettering Parkway, Kettering, NN15 6XR; JT Rehab Ltd, Sheffield, UK

**Keywords:** Strength Training, In-patient care, Deconditioning, Rehabilitation

## Abstract

**Background:** Deconditioning due to in-patients’ stays is predictive of delayed discharges and readmissions; preserving muscle mass and strength in order for patients to remain independent should be of utmost priority. Progressive resistance training offers the most cost-effective way of doing this; however, it is not routinely done.

**Objective:** To examine the feasibility, usability and acceptability of a novel in-bed strength training device (S-Press) by patients and staff members within an in-patient rehabilitation ward in the NHS.

**Methods:** Using a mixed methods approach, 6 patients who performed resistance training on the S-Press device and, alongside 3 healthcare professionals, were interviewed. Data were thematically analysed to examine individuals’ perspectives and perceptions on the feasibility, useability and acceptability of the S-Press. In addition, measures of sit to stand and muscle ultrasound were conducted before use and before discharge. For indications of safety, heart rate and blood pressure measures were taken during each session.

**Results:** Patients found the S-Press easy and comfortable to use and enjoyed using it as an adjunct therapy during in-patient stay. Healthcare professionals using the S-Press noted that it improved mobility allowing patients to gradually build up muscle strength in a safe and motivating way. All participants improved sit to stand performance with use of the S-Press. There were no significant differences observed in heart rate or blood pressure during each session, indicating the safety of S-Press use.

**Conclusion:** From this proof-of-concept study, use of the S-Press to improve muscle strength and physiological function in in-patients is feasible and an acceptable intervention. Further work should focus on use with wider patient demographics and conditions.

## Introduction

Prevention of hospital associated deconditioning is an immediate healthcare unmet need, with high risk of its occurrence, given that over 7 million older adults (65+) were admitted into UK hospitals in the annual period 2020-2021 (“NHS Digital, 2021” n.d.). Muscle deconditioning begins within 2 days, with baseline strength reducing by up to 20% in 1 week; 68% of patients are discharged below their pre-admission level of functioning (Kortebein et al. 2008; Falvey, Mangione, and Stevens-Lapsley 2015). These patients are three times more likely to be re-admitted within 30 days and de-conditioning is responsible for 47% of delayed discharges (Lim et al. 2006).

Hospital ‘exit’ is blocked more than ever, with an increasing older population with multiple pathologies, long-term conditions, poor social care availability, and with Intermediate Care (IC) capacity able to meet only half the demand (The King’s Fund, 2018). High bed occupancy levels, and more complex patients cause delays in discharging patients, which has contributed to increases in people having to wait 12 plus hours in A&E for admission to hospital (The Kings Fund, 2018). Deconditioning and slow physical recovery is the cause of 47% of delayed discharges (Lim, 2006).National Audit Office (NAO) calculate the cost to the NHS for delayed discharging of older patients from hospital to be around £800 million a year, with a negative impact on patient health and outcomes (House of Commons Committee of Public Accounts 2016).

Preserving muscle strength and function has many contributing factors; with physiotherapy having a key role. Progressive resistance exercise (PRE) is known to increase muscle strength in even the frailest individuals (Suetta et al. 2007) and researchers suggest that PRE should be used more in hospitals to prevent deconditioning and improve outcomes (Falvey, Mangione, and Stevens-Lapsley 2015). There is limited PRE equipment available for physiotherapists, its use compounded by a lack of clinical staff numbers and time available. A new S-Press portable leg strengthening device has been designed by a physiotherapist to improve availability and access to effective PRE for vulnerable older patients, offering the option to support use by two patients at a time, with some patients able to use unsupervised once appropriately set-up, and an overall ease to workload for physiotherapists.

PRE benefits have been well documented since the 1950s, yet physiotherapists working in hospitals rarely use it with patients (Suetta et al. 2007; Falvey, Mangione, and Stevens-Lapsley 2015). There are many reasons for this including health barriers, institutional barriers, changing demand on physios within the NHS and lack of appropriate equipment for effective implementation (Brown et al. 2007; So and Pierluissi 2012; Parker, Sricharoenchai, and Needham 2013). Guthrie Smith (Smith 1943) invented the revolutionary sling/spring suspension format used for decades by physiotherapists. The S-Press utilises this tried and tested principle in a re-imagined /re-designed mechanical gym-equipment style, for portability, accessibility, and ease of use by vulnerable temporarily immobile patients in need of muscle strengthening, even if confined to a bed or chair.

Leg muscle weakness, especially knee-extensor strength, is associated with increased falls risk (Tinetti, Speechley, and Ginter 1988; Wolfson et al. 1995) and decrease in function (Van Roie et al. 2011). Leg strength gain is associated with improvements in sit-to-stand, gait, transfers, stair climbing and confidence (Chandler et al. 1998). There is, however, very limited PRE equipment available for use in hospitals. The development of protocols and medical devices to help exercise and maintain muscle mass and aerobic fitness within a hospital setting should be the first line treatment option (Greysen and Patel 2018) and is paramount to preventing this acute sarcopenia (Welch et al. 2018; Supriya et al. 2021). Indeed, a recent study by (Martínez-Velilla et al. 2019) demonstrated that even a short period of low intensity resistance training (5 days) in a hospital setting was sufficient for 60% of patients to improve their short physical performance battery score; however, this involved structural changes within a ward setting, including creating space and resource for resistance training equipment.

There are many barriers to effective physiotherapy and in-patient rehabilitation. Many patients are often too fatigued, dependent, weak, unwell, unstable, anxious, in too much pain, or have pressure sores, preventing them from participating in effective rehabilitation, standing, and walking exercises (Brown et al. 2007; So and Pierluissi 2012). There is often a lack of space near to the bed area, and dedicated gym rooms (especially since COVID-19) have been closed and commandeered as offices. Large static equipment is expensive, may need mains electricity and is difficult to manoeuvre and store. From our own public-patient involvement in research there are a number of barriers associated with trying to increase physical activity within a hospital setting; these include lack of space, lack of equipment and resources and lack of time, all of which the S-Press device has been designed to remedy.

The aim of the study was to assess the feasibility, usability, acceptability and efficacy of use of the S-Press by NHS staff and in-patients. The secondary objectives were to quantify acute and longer-term physiological adaptations in using the S-Press device in a hospital setting that underpin improvements in physical function.

## Methods

### Study Overview

Using a single-arm, mixed methods study design, this proof-of-concept study was conducted during the COVID-19 pandemic in an inpatient rehabilitation ward in a NHS Trust, UK as a real-world evaluation. The proof-of-concept study was approved by NHS ethics (health research authority [HRA] REC reference: 20/YH/0115) and studies were conducted in line with the Declaration of Helsinki. A total of 36 sessions were conducted in eight complex patients with multimorbidity, six patients completed the study, with two participants withdrawing (one self-discharged following 101-day hospital stay, and one was unable to complete the leg press exercise due to uncontrolled pain (from neck of femur surgery) before completing post assessments, one further patient did not participate in the qualitative interview. The study was registered retrospectively with clinicaltrials.gov (NCT06175728)

#### Sampling

Sampling was non-probabilistic, accessible and purposive (Lynn 2016). Participants were chosen if they had used the S-Press as a patient, *n = 6* (quantitative and qualitative data) or as a professional *n* = 3 (qualitative data). Data were collected from one hospital site, where the S-Press was being trialled with appropriate patients.

Inclusion criteria for patients in the study was: any NHS in-patient admitted onto ward site, assessed as medically stable and physically able to participate, and able to provide informed written consent for participation. Exclusion criteria: any patient unable to give informed consent, significantly confused patients, or those unable to do leg press exercises. In addition, there were certain medical exclusions (those who were unstable or with deteriorating medical conditions, acutely unwell with an infection, undergoing treatment for cancer, myocardial infarction within the past six months, or high blood pressure that is uncontrolled). Descriptive information about patient participants is summarised in Table 1.

**Table 1:** Participant Characteristics.

| Participant | Sex | Age Range | Reason for Admission | Rationale for Physiotherapy /Use of S-Press | Number of times used S-Press | Interviewed |
| --- | --- | --- | --- | --- | --- | --- |
| 1 | F | 85-90 | Fractured Femur with intermedullary nail insertion | To teach and assist to walk again properly. | 8 | Yes |
| 2 | M | 70-74 | Neurological issues, Frequent faller | Deconditioning and unable to walk | 6 | Yes |
| 3 | F | 70-74 | Burst Appendix | Deconditioning | 5 | Yes |
| 4 | M | 50-54 | Miller Fisher Syndrome | Deconditioning | 5 | Yes |
| 5 | F | 70-74 | Liver surgery and embolism | Deconditioning, difficulty walking and standing. | 3 | Yes |
| 6 | M | 70-74 | Hip Fracture | Deconditioning, Cognitive Issues | 5 | No |

Three (one male and two female) healthcare professionals (Health Care Assistant (S1), Therapy Assistant Practitioner (S2), Senior Occupational Therapist (S3)), were also recruited (only for the interviews). Inclusion criteria was any NHS staff delivering therapy care to inpatients, and exclusion criteria comprised any staff who do not deliver therapy care (for example catering and domestic staff).

### S-Press Device

The S-Press is a portable, therapeutic level, progressive, leg strengthening device, which can be used on a bed or in a chair. It incorporates between 3kg and 20kg of resistance, across five interchangeable levels, and works bidirectionally so that all the muscles in the legs, important for the sit to stand action, can be strengthened therapeutically and effectively. The use of the S-Press device was incorporated into daily physiotherapy rehabilitation in addition to usual care.

**Figure 1:**
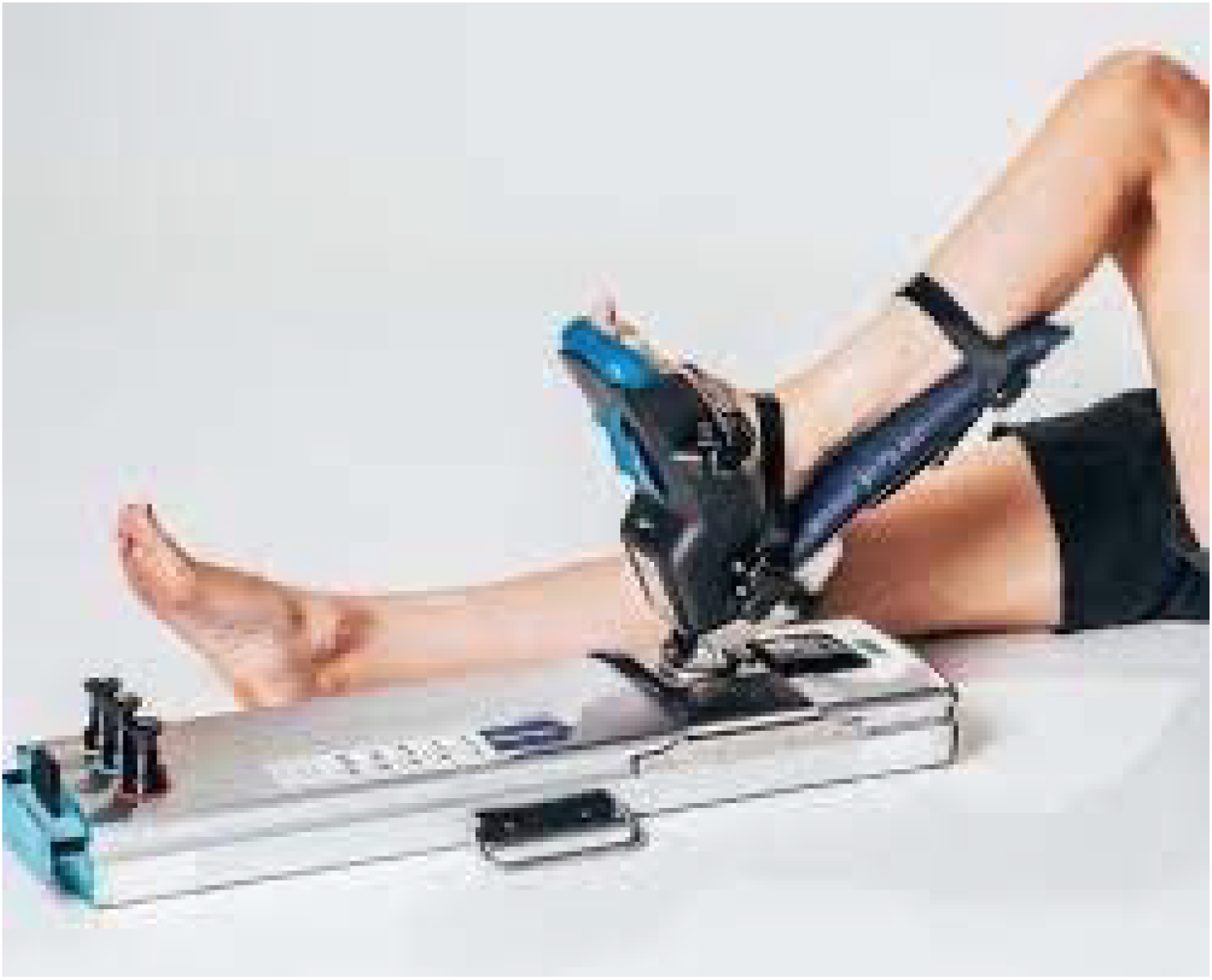
Illustration of the S-Press device

### Exercise Sessions

Staff were trained to set up and deliver the S-Press exercise session with the patients. The exercising undertaken with these patients was during their normal 30-minute physio session. Initial assessment of appropriate resistance level was assessed at the beginning of the first session, by examining the patient participants’ repetition max (1RM) level (maximum resistance level ability). Patient participants were asked to complete 3 sets of 10 repetitions on each leg firstly for the knee extensors and then the knee flexors. A two-minute rest was allowed in between sets and legs, although more rest was given if required by the participants. Blood pressure and heart rate was measured prior to the exercise session commencement, mid-way and at the end of the exercise session, by hospital staff, to ensure that there was maintenance of blood pressure and heart rate and to monitor any adverse effects. The aim was to implement the intervention for six weeks, offering two sessions per week. This was not always practical, as some patients were discharged early.

### Physiological Measurements

Physiological and functional assessments were conducted at baseline (start) and shortly before discharge. Post testing was conducted approximately 48 hours post final exercise session to allow for any fluid changes within the muscle to subside and minimise fatigue of participants.

#### Familiarisation

Following obtaining informed consent, participants were provided with an opportunity to have practice attempt/s at each of the assessments and also the exercise session. For the exercise sessions this was used to identify the starting point in terms of resistance for each individual. All exercise sessions and assessments were performed on the ward

#### Muscle thickness

The muscle thickness of the Vastus Lateralis of each leg was determined by Handheld B-Mode Ultrasonography (Probe: 46mm, 6-12Hz; Orca Medical, Healcerion, UK). The measurements were conducted at 50% of femur length, determined by the distance between knee joint space and greater trochanter as defined by palpation (Balshaw et al., 2019). An experienced blinded investigator analysed images for muscle thickness using ImageJ Software (National Institute of Health, USA) defined as the mean distance between deep and superficial aponeurosis at the proximal, mid-point and distal parts of the image. Image quality for 1 participant was insufficient for analysis due to subcutaneous fat depth.

#### Five times sit to stand

Participants were timed for how long it took them to complete five times to stand. Participants were allowed to use their ambulatory aides if required and this was noted. Where participants couldn’t manage 5 repetitions, the number of complete movements and the time it took for those were recorded. One participant was unable to complete any sit to stand at both pre and post assessment.

### Qualitative Methods

One-to-one semi-structured interviews were undertaken with patients and healthcare professionals to examine their perspectives and perceptions on the S-Press feasibility, useability and acceptability. For this exploratory examination, as informed by Rothstein et al. 2018, these factors were defined in the following way: *Feasibility* was if and how application and implementation of the S-Press was achieved, accounting for how embedding it in routine practice could be achieved. *Useability* was if the S-Press functioned in the way it should, and if it enhanced outcomes and aided physio input. *Acceptability* was if the service users and professionals liked the device, how it worked and its design.

#### Qualitative Data analysis

Qualitative Thematic Analysis (TA) was undertaken, as informed by (Braun and Clarke 2006) involving a six-step framework analysis approach. Interviews were coded using a priori topical codes according to the three assessment areas of feasibility, usability, and acceptability. NVivo was used to organise and analyse the data. The six-steps followed comprised: (i) familiarisation with the data (data were transcribed, read, and re-read, and initial codes noted); (ii) generating initial codes (interesting features across the data set were coded, and data relevant to each code collated); (iii) searching for themes (codes were collated into potential themes, and data relevant to them gathered); (iv) reviewing themes (themes were assessed, combined, refined, separated, and discarded accordingly); (v) defining and naming themes (operationalisation of each theme and subthemes and development of clear working definitions that capture the essence of each theme); and (vi) producing the report (the overall learning from the qualitative data analysis was written up, summarising the themes and illustrating these with noteworthy quotes). This process was completed with attention to the guidance on principles of credibility, transferability, dependability and confirmability (Korstjens and Moser 2018), following guidelines by (Shenton 2004).

## Results

### Physiological Data

There was no change in mean heart rate using the S-Press (Pre: 82 ± 13 b.min^-1^; Mid: 84 ± 15 b.min^-1^ and Post: 84 ± 15 b.min^-1^) nor blood pressure (Pre: 125 ± 15/ 76 ± 8 mmHg; Mid: 122 ± 13/ 75 ± 8 mmHg and Post: 124 ± 14/ 77 ± 8 mmHg). There were no reported adverse events during or after use of the S-Press.

### Sit to Stand performance

From baseline measurements of sit to stand performance, all six participants were classified as pre-sarcopenic according to EWGSOP guidelines. At discharge, two out of the six participants were no longer classified as pre-sarcopenic (5STS <15 seconds). The participants able to complete sit to stands all improved five times sit to stand performance following use of the S-Press with mean five times sit to stand performance (Pre: 35.7 ± 13.8 seconds vs Post: 19.6 ± 12 seconds). At Pre-assessment, one participant was only able to perform two sit to stands in 52.1 seconds, at post assessment the participant was able to perform all 5 sit to stands at discharge in 18.2 seconds. 1 participant was unable to perform any sit to stand at pre or post measurements, this patient had had surgery to insert a stent due to a previous glioblastoma diagnosis, and was unable to manage a safe sit to stand. There was a mean increase of 15.4 mm (17%) in vastus lateralis muscle thickness following s-press intervention.

### Qualitative Results

The emergent themes associated with each of the three assessment areas of ‘feasibility’, ‘usability’, and ‘acceptability’ are presented in Figure 3. The assessment areas yielded a range of individual but interlinked themes, which were identified from the narratives of the patients and staff.

**Figure 2:**
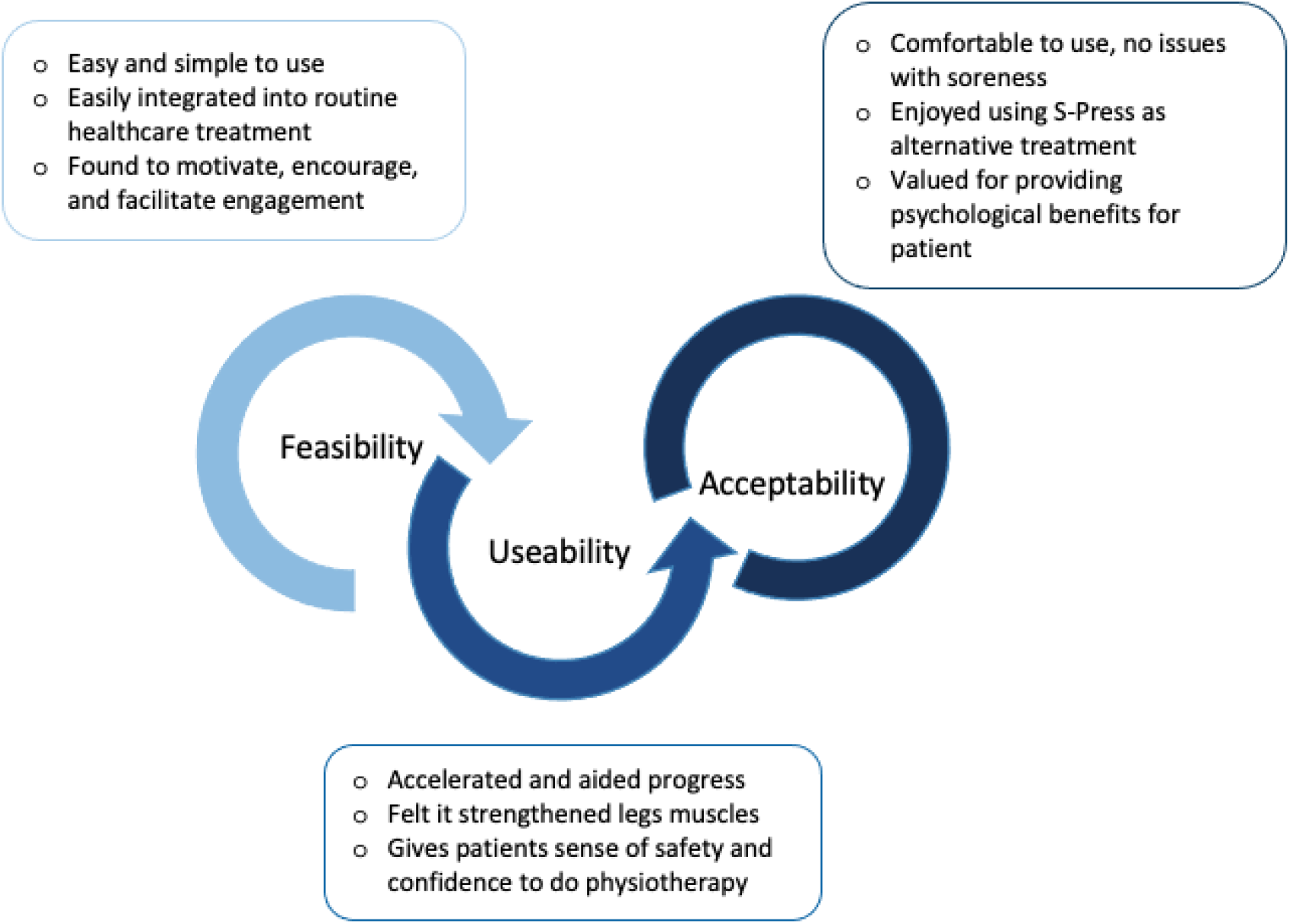
Summary of key findings for feasibility, useability and acceptability subthemes from semi-structured interviews.

### Feasibility

This assessment area was made up of three themes around if and how application and implementation of the S-Press was achieved, accounting for how embedding it in routine practice was accomplished.

### Easy and simple to use

Across all the narratives, both from a patient and staff perspective, the S-Press was easy to use and implement. The patients found that when taught how to use it, application and implementation was easy.

> **P2:** *Well, you’ve just got to put your leg in the machine and keep pumping and then after eight pumps you give them two really good, hard pumps, make sure your legs are alright. Yes, that was alright. I had no problem. I found it helpful that someone taught me how to use it, but as soon as I was taught, it was really easy to use. Yeah, it was easy*.

The staff also identified that it was easy to teach the patients how to use the S-Press; this was done through verbal instruction and actively doing it. As the patients worked with the staff, they learned quickly how to use it with no one experiencing any difficulty.

> **S3:** *They all seem to respond quite well to just verbal direction, and you’re actually actively doing it as well. We had a really good response from our patients in terms of sort of picking it up quite quickly, there was nobody that it was difficult with*.

A key factor identified was that it was not a complicated piece of equipment and could be easily embedded within routine practice. The patients referred to the simplicity of it highlighting that it was easily done and engaged with.

> **P3:** *Well, I’m strapped into it, and I just had to push. Which is simple. Just a matter of pushing your legs. Well, one at a time. There’s just nothing complicated about it. Nothing complicated at all about it. I found it so easy to use. And that was a good thing from my point of view. You know, something simple to get yourself going*.

One factor relevant to staff, was they found it easy to clean, so they could use it confidently with different patients, knowing that infection control could be addressed, and was not problematic.

> **S1:** *So, we have some cleansing wipes which are on the ward anyway, so we just wiped down the metal holder for S-Press, the footplate, just wiped it completely the whole way through. It then went back into the storeroom and where we kept it and no issues at all, with the cleaning. It was easy to clean. There were no difficulties*.

### Easily integrated into routine healthcare treatment

The S-Press was simple to incorporate into the patient’s care. Both the patients and staff identified it was not onerous in terms of time. This was something staff felt helped with engagement meaning it was effective and straightforward to embed in the patient’s day.

> **S1:** *And it’s quite a straightforward thing you can do, depending on the patient, and how long it takes them to do a certain number of reps. You can do it within 10 or 15 minutes, it’s quite easy. It’s quite easy to spread out through your day. So, then you’d be a lot more willing to engage*.

The patients felt that the S-Press was an addition to their care that offered them more opportunity to exercise and do physiotherapy than they might ordinarily have experienced. It was easy to use, it didn’t hurt them, with patients finding they could complete all their sessions.

> **P1:** *Yes, it definitely offered me the opportunity to exercise more than otherwise. They came and I was able to use it regularly, and because it was easy to use, and didn’t hurt, and seemed to work, I did all the sessions*.

The S-Press also gives an alternative form of exercise for some patients. The device was something that could be adopted as part of their rehabilitation

> **S3:** *There’s only so much standing practice you can do… the S-Press, it’s a real targeted those muscles that they need to stand, without the need to actually stand because they may get half a stand and be completely exhausted, whereas the S-Press, if they’re resting on the bed, you can adapt, you’ve got a good range of resistance that you can use, you can build it up*.

### Found to motivate, encourage and promote engagement

The patients found that they were motivated to use and engage with the S-Press, which all facilitated the implementation. Patients felt gains from using it, that they were building strength and progressing, and were actively doing something to aid their recovery. This seemed to motivate patients to want to use the S-Press and carry on with the sessions.

> **S2:** *I think because he could see that he was progressing. I think they like the fact that…it’s helping them, if they can see an improvement, that’s when they like to use it. You know, are you bringing it this week? Or, you know, when’s my next session? You know, get some people who are really keen*.

Patients were particularly motivated by the increase in resistance that they could aim towards as they continued to use the device. This provided feedback for the patients, they were encouraged to continue engaging with the S-Press, working towards the higher resistance.

> **P4:** *It was quite good having the different numbers of something to work towards, as that kind of motivates you. I mean, I’ve got to five pretty quick on my right leg, and then I was on two to start with on my left leg, and I think we got up to four, within five sessions. So, if we were still doing that, probably still, I’ll probably be on five on my left leg now, definitely*.

The S-Press was used for goal setting for the patients, which motivated them further. These were targeted goals, which were achievable, and helped patients psychologically as they could monitor this and see they were progressing.

> **S1**: *I suppose it gave them small, targeted goals, which they can target on, in general, but on the S-Press, or I want to try and do at least one set at the next level, and then they could give them small, achievable goals, which helped them psychologically, that I’m improving*.

A final element was the interaction with the staff and the feedback given by them; the S-Press and these interactions were found to encourage and motivate the patients and get them to engage, which aided the implementation and feasibility.

> **P1:** *And, you know, with them being with me, guiding me and saying how I was doing. I found it all very helpful. The social aspect of it helped, working with somebody, and having them support me. That encouraged me and motivated me, which was really good*.

### Useability

When examining useability, five themes were found across the data set, which were all related to if the S-Press functioned in the way it should, if and how it enhanced outcomes and if and aided physiotherapist input.

### Accelerated and aided progress

Participants discussed the ways that they felt using the S-Press aided their progress and accelerated their recovery. One of the participants compared this way of doing physiotherapy with other previous treatments, finding they progressed quicker with the S-Press.

> **P1:** *It* [S-Press] worked *very well. I could bend my knee better and much more quickly using that than I did when I had my knee done, at the same time. I learnt to bend my knee and straighten my leg much quicker than I did when I had physio when I had my new knee. I found this was a much quicker way of moving*.

The participants also talked about how they felt they would not have progressed their mobility if they had not used the S-Press. They discussed different ways in which they progressed such as getting them home quicker:

> **P3:** *I felt happy. I could build up my muscles and get home. It was something that could really help me get home quicker, get home sooner*. walking without aids:

> **P4:** *I don’t think I’ll be where I am now, without the crutches. I’ll think that, you know, the S-Press has helped me get the muscles back working, definitely. So yeah, you can put that down, I probably, I would probably still be relying on my crutches*.

and mobility and movement (which again speeded up discharge):

> **S1:** *I think on the whole it tended to, the ones, the ones who were we saw quite a bit of progress with the S-Press got them mobile and got them moving quicker, which allowed us to start planning to get them home quicker*.

### Felt it strengthened legs muscles

All five patients interviewed vocalised that they could feel strengthening of their muscles and legs: *I felt that I’d got more strength in my leg. That the feeling that my muscles were being built up* (P1); *I think it strengthening my leg, it felt like I got strengthen in my leg. I feel I am getting a stronger leg and muscles* (P2); *I feel as if my muscles have got stronger* (P3); *I could really feel that my legs and muscles were being helped* (P4); and *It started to strengthen the muscles above my knees* (P5).

The staff also noted that the patients experienced strengthening of their legs, and how this enabled them to achieve positive outcomes in a way that was not difficult or taxing for the patient. It was felt that there was a notable difference for the patient from before to after using the S-Press.

> **S1:** *One of the men we saw, he couldn’t stand beforehand. He didn’t have the strength in his legs to straighten his legs to stand up. But we got him to the point he could stand up for 30 seconds. So, he could feel the strength, he was improving, and he felt like his legs, it wasn’t as taxing to try and do it*.

It was suggested by one of the patients that the S-Press was a better way of improving their muscle strength than the more traditional type of physiotherapy input, indicating that this was a preferred and quicker way to aid them.

> **P3:** *It got, it helped me work my legs quicker, getting them stronger than having to wait and then just doing the physio walking up and down and walking up steps. The actual pushing strengthens the muscles better*.

### Gives patients sense of safety and confidence to do physiotherapy

The S-Press gave the patients confidence to do their exercises and their physiotherapy. They had confidence in the S-Press, a belief that it was helping and would work, and in turn confidence in themselves and what they could do and achieve.

> **P1:** *Well, I did have confidence in it. I think it gave me confidence; you know what they were doing. I mean I am quite fit normally, anyway. And I felt this was helping me get back to my fitness…It gave me confidence to try and walk and you know move*.

The S-Press was found to give the patients confidence to progress gradually with their physiotherapy, their mobility and movement and building the strength in their legs.

> **S1:** *S-Press and it just helped her to gradually progress from standing from the lower chairs to being able to stand up from the raised toilet seat and frames being able to walk down the corridor, just helped with her confidence and helped build up the strength in her legs*.

Both staff and patients suggested that psychologically the S-Press gave them confidence. Using it they could safely push themselves and undertake the exercises. This gives patients confidence to just ‘get on and do’ the exercises they need to do; this would ultimately help them and aid their recovery and progress.

> **P3:** *It built my strength up from a physical point of view, but also mentally. Mentally and psychologically, it gave me confidence to push myself. And sitting down you are also more confident that you are not going to fall or something. It’s given me confidence to get on and do all the exercises I need to do*.

### Acceptability

The final element of the analysis examined the acceptability of the S-Press and if the patients and professionals liked the device and its design, comprising four themes.

### Comfortable to use, no issues with soreness

All the patients commented that they liked the design in relation to comfort when using and having physiotherapy with S-Press. They all expressed there was no soreness, rubbing, or skin irritations experienced. They also all commented how they felt the S-Press was well padded which added to the comfort and to the avoidance of soreness. Common discussions included:

> **P1**: *No none at all, nothing. It was well padded and didn’t cause any soreness*.

> **P3:** *I was fine with it. I can’t think of another way. I mean, it’s all padded. So, nothing rubbed you. The straps weren’t, the straps weren’t too tight. They fitted comfortably around my legs and feet*.

> **P5:** *No, not at all. I got no soreness anywhere on my legs from the S-Press*.

The staff agreed that the S-Press appeared to be comfortable for the patients and did not cause issues in relation to skin problems, soreness, or pain. They commented on how the straps were all adjustable and so could be adapted for each patient to enhance the comfort.

> **S3:** *No, we didn’t, we didn’t have any issues at all. It was very padded. And yeah, there’s the straps where it could be sort of tightened and loosened, so they didn’t particularly dig in or we didn’t have any issues with the strap*.

Staff also found that there were no issues of skin viability, and with care they could work around problematic areas that patients may have with their skin. They discussed how they could put things in place to protect the patient and maintain the comfort and safety of using the device.

> **S1:** *If they needed leg dressings, we obviously wouldn’t recommend to take the leg dressings off before, but you could quite easily put a pillowcase between them…. There wasn’t any issues of skin viability*.

### Enjoyed using S-Press as alternative treatment

Patients reported they enjoyed using the S-Press as part of their physiotherapy and this enhanced its useability: ‘*I did really enjoy exercising with it’* (P1); ‘*I suppose I enjoyed it all really because it made me better’* (P2); and ‘*Very, very beneficial, and enjoyable’* (P5). One of the patients explained how they looked forward to having the sessions, finding them all beneficial and enjoyable.

> **P4:** *I was like I said, I was like, literally looking forward when, I didn’t know when she, when she was gonna, come for the sessions. When she came in, I was like, yeah, okay, brilliant, let’s crack on…very, very beneficial, and enjoyable*.

The staff also commented how they felt that the patients enjoyed using and engaging with the S-Press, with some patients asking to continue to use it after they had completed all their sessions. In addition, one of the staff noted how they enjoyed doing this in comparison to other types of physiotherapy sessions, as they liked the way the machine worked offering resistance, finding other exercises without the S-Press tedious.

> **S1:** *The ones who really enjoyed doing, felt it was different to what the other exercises are doing. They really liked the resistance element and that they had to work a bit harder from different levels to progress. They enjoy it as it gives you an alternative to trying to do exercises in a bed which can be a bit tedious at times*.

Both the patients and staff commented that the S-Press offered a social interaction and one-to-one targeted care and rehabilitation. This added to the enjoyment of using the S-Press, and in turn the wellbeing and recovery for the patients.

> **S3:** *I guess, an intensive interaction, the therapies, they would certainly have known they had a dedicated therapist for them at that time. So, I think the wellbeing element was probably the investment of staff in their rehab, and the patients enjoyed the time with the therapists working with them*.

> **P1:** *I did really enjoy exercising with it. It became more of a social thing really. I was in a room on my own. So, it was, I quite enjoyed it when they came*.

### Valued for providing psychological benefits for patients

This theme is about the acceptability of the device as the S-Press was valued as a way of giving the patients a psychological benefit. The device therefore not only works on the physical recovery for the patients, but it also seems to support and assist with their mental wellbeing.

> **P4:** *So, when I went to rehab, it was sort of like, great, and then when I signed up for the S-Press, it was like, now I’m on the last leg for going home. It was all, all like a boost for me really. My mental wellbeing. I keep repeating myself, but it was the start of the process to get back to get back to some sort of normality*.

The staff also commented how there was both physical and psychological value gained from the device, whereby the physical gains that were experienced, were associated with a positive psychological impact.

> **S1:** I think it was an element of physical and psychological because physically, he can see his legs were strong again, getting stronger. On the whole, it helped, made, give them a brighter mood and positive mood. There was some element of it being psychological that they felt they were doing like a strengthening exercise.

The patients discussed how the S-Press was psychologically beneficial and this meant that they had a belief that they would get better, meaning it encouraged them to engage with and use the device.

> **P2**: *It was kind of, of mentally beneficial. You are hoping that you are going to get better, it just makes it, you know you feel better by doing it I was just hoping it would make me better, that’s the main thig, it helped me mentally thinking I would get better. And so I just keep going - do this do that and I done it. It did really help*.

## Discussion

The aim of the study was to assess the feasibility, usability, acceptability and efficacy of use by NHS staff and in-patients of the S-Press (a novel progressive resistance exercise machine) in a hospital bed setting. The secondary objectives were to quantify acute and longer-term physiological adaptations in using the S-Press device in a hospital setting that underpin improvements in physical function.

For use of the S-Press, each patient session using the device took less than 10 minutes to perform, with patients and staff finding it easy to use and include within rehabilitation programmes. Staff were able to verbally instruct patients on how to use the device comfortably and adequately perform appropriate cleaning and infection control measures. Patients in particular felt that the incorporation of PRE within their rehabilitation programmes was more beneficial than other methods and provided an ideal opportunity to exercise safely throughout the day. The S-Press was shown to be comfortable, safe, and easy to use by all participants, with no negative implications or adverse events reported.

Patients showed a great enthusiasm for participation and use of the S-Press. Patients felt empowered and more in control of their recovery. They could see a clear objective improvement with every session in their leg muscle strength, through being able to manage the next resistance level up. They had more confidence in their physical abilities, which carried over into their other therapy interventions. They felt they were improving quicker than they would have without it or compared to previous hospital admissions. Many wanted to do more sessions per week with it and to take the S-Press home with them. All recommended the use of the S-Press for rehabilitation.

This patient experience and satisfaction is an important indicator of rehabilitation success and clinical effectiveness (Eversole et al. 2021). Improving patient experience is not only good for patients but associated with better clinical processes and outcomes (Doyle, Lennox, and Bell 2013; Kingsley and Patel 2017); however, this may be inherently biased (Beattie et al. 2015; Medina-Mirapeix et al. 2015; Eversole et al. 2021) especially where patients perceive they have had better treatment than others (Beattie et al. 2015). That said, high adherence within an inpatient setting (such as the current study) and higher self-efficacy have been demonstrated to improve overall treatment adherence (Jack et al. 2010).

A recent study by (Coleman et al. 2021) demonstrated that PRE was safe and acceptable intervention in a similar patient group to the current study, likewise reporting no adverse events during the trial, which is also in line with other studies (Martínez-Velilla et al. 2019; Tibaek et al. 2017; Coleman et al. 2021). In the current study, the physiological response to using the S-Press did not differ from that at rest, with only a small increase in heart rate and blood pressure following acute use of the S-Press, demonstrating safety of using the device in a range of patient conditions. This physiological response is often a limiting factor to prescribing resistance exercise within clinical patient groups often reporting large increases in blood pressure in acute settings (MacDougall et al. 1985) despite an overall reduction in blood pressure often observed over resistance training interventions (Cornelissen et al. 2011).

Previous studies in older post-acute inpatient populations have observed improvements in muscle strength and function following bed based PRE (Mallery et al. 2003; Kawakami et al. 2001) and chair-based PRE (Latham et al. 2003; Suetta et al. 2007; Coleman et al. 2021), in line with the findings from the current study. In the current study, we observed an improvement in the participants’ abilities and speed of sit to stand accounting for a mean 52% improvement to perform five sit to stands, and an average of 18.41 seconds faster for five times sit to stand. Importantly at discharge, two out of the six participants were no longer classified as probable sarcopenic according to the EWGSOP guidelines (Cruz-Jentoft et al. 2019) representing a lower risk of loss of independence, falls, readmission to hospital and mortality. All participants in the current study improved their ability to perform sit to stands.

Deconditioning is a feature of long inpatient stays with substantial loss of muscle mass during periods of ambulatory care. In the current study, whilst not statistically powered to determine efficacy for improvements in muscle mass we demonstrated an objective stability or improvement in muscle thickness of all participants using b-mode ultrasound.

## Limitations

This is a first in man study using the S-Press device and, whilst our primary outcomes were to look at usability and feasibility of the S-Press in an NHS in-patient hospital setting with staff and patients, we are statistically underpowered to infer physiological adaptations attributable to the use of the S-Press. The chosen ward for the current project had a low patient turnover rate which may have limited recruitment. In addition, this investigation was conducted during COVID-19 lockdown periods in the UK which affected the ability to conduct the trial sufficiently within an NHS setting due to staffing illness and other governance constraints around conducting research during this period. Our patient group was heterogeneous with a mix of neurological, orthopaedic and surgical interventions and, whilst this could be considered a strength of the research in demonstrating the usability of the S-Press across patient groups, a more robust homogeneous patient group should be considered in further research as should a usual care control group.

## Conclusions

The S-Press device provides a feasible, safe and acceptable modality to perform progressive resistance exercise in an inpatient population within an NHS rehabilitation programme. Patients enjoyed the use of S-Press and felt that it accelerated their recovery and subjectively improved their physiological and psychological recovery following admission. This was coupled with large improvements in physical function as assessed by the five times sit to stand test. Further robust research is required to show efficacy of the S-Press device for improving physical function in this patient group.

## Data Availability

All data produced in the present study are available upon reasonable request to the authors

## Funding details

This work was supported by the Versus Arthritis UK under Grant (number 22006) The S-Press device and JT Rehab Ltd have been awarded funding from the European Regional Development Fund through the Sheffield Innovation programme, Advanced Wellbeing Research Centre Accelerator Funding (Research England), Academic Health Science Network East Midlands and Versus Arthritis UK, Catapult funding from the European Regional Development Fund and Innovate UK Women in Innovation.

## Disclosure statement

JT is owner of JT Rehab Ltd who are inventors and creators of the S-Press device that has a direct application of this research. No other authors have competing interests to declare.

## Notes

### Clinical Trial

NCT06175728

### Author Declarations

The proof-of-concept study was approved by NHS ethics (health research authority [HRA] REC reference: 20/YH/0115)

